# Insulin Resistance’s Impact on Cognition in Middle Aged Adults from the PREVENT cohort: Interactive Effects with Depression

**DOI:** 10.1101/2023.01.31.23285252

**Authors:** Sarah Bauermeister, Graham Reid, Michael Ben Yehuda, Gregory Howgego, Karen Ritchie, Tam Watermeyer, Sarah Gregory, Graciela Muniz Terrera, Ivan Koychev

**Affiliations:** Department of Psychiatry, University of Oxford, Oxford, OX37J; Edinburgh Dementia Prevention, Centre for Clinical Brain Sciences, University of Edinburgh, Edinburgh, EH4 2XU; Institut de Neurosciences de Montpellier, Inserm, Montpellier, France

## Abstract

**Background:** Alzheimer’s disease, type 2 diabetes mellitus, and depression are significant challenges facing public health. Research has demonstrated common comorbidities amongst these three conditions, typically focusing on two of them at a time.

**Objectives:** The goal of this study, however, was to assess the interrelationships between the three conditions, focusing on mid-life risk before the emergence of dementia caused by Alzheimer’s Disease.

**Methods:** In the current study, we used data from 665 participants from the prospective cohort study, PREVENT.

**Findings:** Using structural equation modelling, we showed that (i) insulin resistance predicts executive dysfunction in older but not younger adults in midlife, that (ii) insulin resistance predicts self-reported depression in both older and younger middle-aged adults, and that (iii) depression predicts deficits in visuospatial memory in older but not younger adults in midlife.

**Conclusions:** Together, we demonstrate the interrelations between three common non-communicable diseases in middle-aged adults.

**Clinical Implications:** We emphasise the need for combined interventions and the utilisation of resources to help adults in midlife to modify risk factors for cognitive impairment, such as depression and diabetes.

**Funding:** The PREVENT study was funded by the Alzheimer’s Society (grant numbers 178 and 264), the Alzheimer’s Association (grant number TriBEKa-17-519007) and philanthropic donations. GR acknowledges funding for this work for his research programme funded by the Medical Research council (Dementias Platform UK) and Five Lives Ltd. IK declares funding for this project through Medical Research Council (Dementias Platform UK), NIHR Oxford Health Biomedical Research Centre and NIHR personal awards. SG acknowledges funding for salary from the Medical Research Council Nutrition Research Partnership Collaboration Award (MR/T001852/1).

**Key Messages:** *What is already known on this topic:* Mood disorders and metabolic diseases are known to be frequently comorbid. Furthermore, both conditions are known to be associated with cognitive impairment and cognitive decline. There has been some evidence that the risk of cognitive impairment associated with diabetes and depression is most notable in midlife. However, studies focusing on this period of life have been sparse and most research has modelled bivariate correlations amongst cognitive impairment, depression, and diabetes. As such, this study was conducted in order to model the interrelations between the three conditions in a large cohort, whilst focusing on midlife as depression and diabetes in this period are thought to carry higher risk for cognitive impairment.

*What this study adds:* Whilst insulin resistance, as a core feature of diabetes, was related to depression across all stages of midlife, the relationship with cognitive functioning was more complex. In the current study, we found that the stage of midlife in which middle-aged adults find themselves moderates the relationship between insulin resistance and cognition and depression and cognition. That is, only in older middle aged adults does insulin resistance predict impaired cognition (i.e., executive function) and does depression predict impaired cognition (i.e., visuospatial memory).

*How this study might affect research, practice or policy:* Clinicians should be mindful of the impact of comorbidities between cognitive impairment, metabolic diseases, such as diabetes, and mood disorders, such as depression in midlife. Given the risk of intractable dementia in individuals with cognitive impairment, available resources for intervening in modifiable risk factors, such as depression and diabetes, should be considered for adults in the middle period of life.

## 1. Introduction

The prevalence of Alzheimer’s disease (AD) and type 2 diabetes mellitus (T2DM) is reaching epidemic proportions across the globe. Indeed, numerous studies have shown that those with T2DM are at risk of developing AD and that the AD brain, in turn, becomes even poorer at processing glucose as the disease progresses (Arnold et al., 2018). Central nervous system deficits in glucose processing are defined as central insulin resistance, which typically impacts the brain’s ability to support basic psychological functioning, including cognition and mood (Lee et al., 2016). However, studies have also shown that with comorbid AD, psychological dysregulations associated with brain insulin resistance occur at an enhanced degree (Talbot, 2014). Likewise, healthy adults without a T2DM diagnosis, but with higher levels of insulin resistance, also have higher risk for abnormal cognitive and affective functioning (Reijmer et al., 2010). Longitudinally, those with higher levels of insulin resistance, even in the absence of a T2DM diagnosis, have a higher risk of AD just three years later (Schrijvers et al., 2010). Yet whilst there is clear evidence of a link between insulin resistance and cognitive impairment, there remains a paucity of research exploring related variables, such as affective disorders, which would enhance our understanding of the relationship between AD and T2DM in ageing populations (Bauermeister & Bunce, 2015).

A better understanding of dementia risk factors and their interactions is a priority given the inefficacy of available treatments and the fact that dementia-related brain changes occur decades before the expression of any dementia symptoms (Ojakäär & Koychev, 2021). It is estimated that up to 35% of dementia cases are attributable to preventable risk factors (Livingston et al., 2020). Over the last 10 years there has been increasing interest in the metabolic aspects of AD, focusing mainly on the dysregulation of glucose, as well as some lipid compounds. As for T2DM, research has suggested that a T2DM diagnosis carries a 1.5 times higher risk of non-vascular dementia compared to the general population (Chatterjee et al., 2015). Indeed, patients with AD seem to have reduced peripheral insulin sensitivity and resting hyperinsulinaemia, with evidence that their cognitive function may be improved by inducing further hyperinsulinaemia while maintaining euglycemia (Craft et al., 1996). This suggests a chronic alternation in patients’ metabolic state, leading to cognitive impairment with at least some degree of reversibility as demonstrated by a recent case-control study showing reduction in dementia incidence with diabetic agents that cross the blood-brain barrier (Wium-Andersen et al., 2019). Animal models of both AD and insulin resistance have similar phenotypes in terms of brain insulin handling, receptor expression, and resistance (Akter et al., 2011). A number of mechanisms for the effect of diabetes on cognition and dementia risk have been suggested. For example, there is some evidence that T2DM, a state of peripheral insulin resistance, is associated with increased Aβ deposition (Stanciu et al., 2020); however, insulin may be acting through other mechanisms such as by increasing inflammation, oxidative stress, vascular pathology, or though altered glucose and lipid metabolism, thereby increasing the likelihood of an AD diagnosis in those with T2DM.

As for related comorbidities, the presence of T2DM more than doubles the odds of co-morbid depression and depression worsens the prognosis, mortality, and treatment compliance of diabetic patients (Bădescu et al., 2016). There is also a small but growing body of evidence which suggests diabetes and depression may act additively to increase the risk of dementia (Sullivan et al., 2013). For example, one study has shown that whilst diabetes and depression differentially impact cognitive processes, such as memory and executive function, together they significantly accelerate the general overall rate of decline, especially in 50-64 year olds (Demakakos et al., 2017). With age-related decreases in cognitive functioning, any additional processing burden caused by affective symptoms, along with the effects of poor blood glucose control, may prove detrimental to cognitive processing. Dementia, depression, and T2DM are thus three common non-communicable disorders, which often co-exist, negatively interact with each other and may share pathophysiological mechanisms. Existing evidence suggests mid-life is when preventable risk factors such as depression and poor T2DM control exert the largest effect on dementia risk (Livingston et al., 2020). However, the exact nature of the interaction between insulin resistance, depression and dementia have not been directly characterised in this age group. Thus, in the current study, we used the PREVENT study cohort of 40-59 year-olds to examine the relationship between insulin resistance, cognitive function, and depression in mid-life prior to a dementia diagnosis.

## 2. Materials and Methods

### 2.1 Participants

665 participants aged 40 to 60 years old were included in the current paper from the PREVENT cohort. Participants were all cognitively healthy at recruitment (i.e., no diagnosis of cognitive impairment) as assessed by self-report and formal cognitive testing conducted via interview, as well as the Addenbrooke’s Cognitive Assessment III.

### 2.2 Computerised Cognitive Tasks

Participants completed a neuropsychological assessment from the COGNITO battery. From the battery of assessments, they undertook tasks which included measures of processing speed (reaction time), episodic memory (recall), and phonemic and semantic fluency. Visuospatial orientation was assessed through the 4 Mountains Test.

### 2.3 Insulin Resistance

Insulin and glucose concentrations were determined by analysis of fasting plasma samples obtained at the first study appointment. Insulin resistance was calculated using the Homeostatic Model Assessment for Insulin Resistance (HOMA-IR): fasting plasma glucose (mmol/l) times fasting serum insulin (mU/l) divided by 22.5.

### 2.4 Depressive Symptom Burden

The level of affective symptoms was determined using the Epidemiological Studies-Depression Scale (CES-D). CES-D is a 20-item measure which asks respondents to indicate the extent to which they have experienced various symptoms of depression, scoring each item from 0 (rarely/none) to 3 (most of the time). Scores range from 0-60 with higher scores indicating greater depressive symptom burden.

### 2.5 Statistical Analysis

#### 2.5.1 Pre-processing

All data processing and statistical analyses were performed in Stata SE 16.1.Cognitive tasks were log-transformed to normalise the distribution where appropriate. Insulin resistance was also log-transformed and two extreme outliers were trimmed for this variable (> 99.9 percentile).

#### 2.5.2 Structural Equation Model

To assess the direct and indirect effects between insulin resistance, depression and cognition a structural equation model (SEM) was used. The aim of the SEM analysis was to assess the shared mechanistic pathways underlying insulin resistance, depression and cognition in a single model. The aim of the SEM was also to assess the prediction pathways of insulin resistance and depression on selected individual cognitive tasks and of insulin resistance on depression. Executive function was assessed as a latent construct formed from the semantic and phonemic fluency tasks, which were initially z-transformed. A measure of intraindividual standard deviation of reaction time was computed from the processing speed task and was included in the model as a proxy measure of neurocognitive integrity (Bunce & Bauermeister, 2019). Depression was included in the model as the total CES-D score and insulin resistance was entered as the HOMA-IR value. To adjust for the confounding effects of age, education and sex, these were also included in the model as covariates.

## 3. Results

The participant sample had a mean age of 51.20 years (*SD* = 5.44) and 61.65% of the participants were female. All baseline participant characteristics are presented in Table 1.

**Table 1.** Participant Characteristics.

| **Demographics** |  |  |  |
| --- | --- | --- | --- |
| Age |  |  |  |
| Sex | 665 | % female | 61.65 |
| Education (years) | 661 | M(SD) | 16.67(3.39) |
| **Cognition** |  |  |  |
| 4 Mountains | 423 | M(SD) | 10.41(2.34) |
| Processing speed | 645 | M(SD) | 5756.29(1516.21) |
| Phonemic fluency | 645 | M(SD) | 11.32(4.12) |
| Semantic fluency | 644 | M(SD) | 16.40(4.15) |
| Recall | 644 | M(SD) | 6.90(1.47) |
| ISD | 554 | M(SD) | 0.14(0.07) |
| **Affective** |  |  |  |
| CES-D | 659 | M(SD) | 9.26(8.46) |
| **Biomarkers** |  |  |  |
| Insulin resistance | 644 | M(SD) | 60.02(40.85) |

### 3.1 Pairwise Correlations

A pairwise correlation with Bonferroni correction was conducted and showed few significant associations between the cognitive variables of interest (outcome variables) and predictor/mediator variables (insulin resistance and depression). The full correlation table output is presented in the supplementary materials (S1) but to be noted are the significant associations between insulin resistance and semantic verbal fluency (r = -.18, p < .01), between insulin resistance and sex (r = .16, p < .01) where females were coded as zero and between insulin resistance and depression (r = .15, p < .01).

### 3.2 Structural Equation Model

#### 3.2.1 Cognitive Variables

A latent construct for executive function (language) was formed from the two indicator measures of phonemic verbal and semantic verbal fluency. Both indicators are sensitive indicators of cognitive change over time as well as early indicators of mild cognitive impairment (MCI) and dementia (Frankenberg et al., 2021). Although both were correlated with each other (r = .298, p < .001) a covariance relationship was not required in the model (non-multicollinearity) as the level correlation did not affect model estimation. Processing speed was entered into the model as an individual task measured as the average of 12 reaction time trials (ms). The 4 Mountains test score was entered into the model as a measure of spatial memory and a delayed recall of names score was entered into the model as an assessment of delayed recall over time. An intraindividual standard deviation metric of the reaction time task was computed to reflect a proxy measure of neurocognitive integrity. Of note is that it is also a sensitive indicator of cognitive change over time, as well as an early indicator of MCI and dementia (Haynes et al., 2017).

#### 3.2.2 Covariate Variables

To adjust for the confounding effects of age, education and sex, these were entered into the SEM model as covariates. Direct paths were extended between the covariates and the cognitive variables. Age was included into the model as a continuous variable. Sex was coded such that females were zero and males were one and education was included as a continuous variable (number of years).

#### 3.2.3 Predictor Variables

Insulin resistance was included in the model as a value in mg/min and depression was included as participants’ total CES-D score.

#### 3.2.4 SEM Measurement Regression Paths

A direct path was extended from insulin resistance to each of the individual cognitive variables and the executive function latent construct (Figure 1). A direct path was also extended from depression to each of the cognitive variables and to the executive function latent construct to assess the relationship between depression and these variables. To assess mediation by depression on the relationship between insulin and cognition, a direct path was also inserted between insulin resistance and depression.

**Figure 1.**
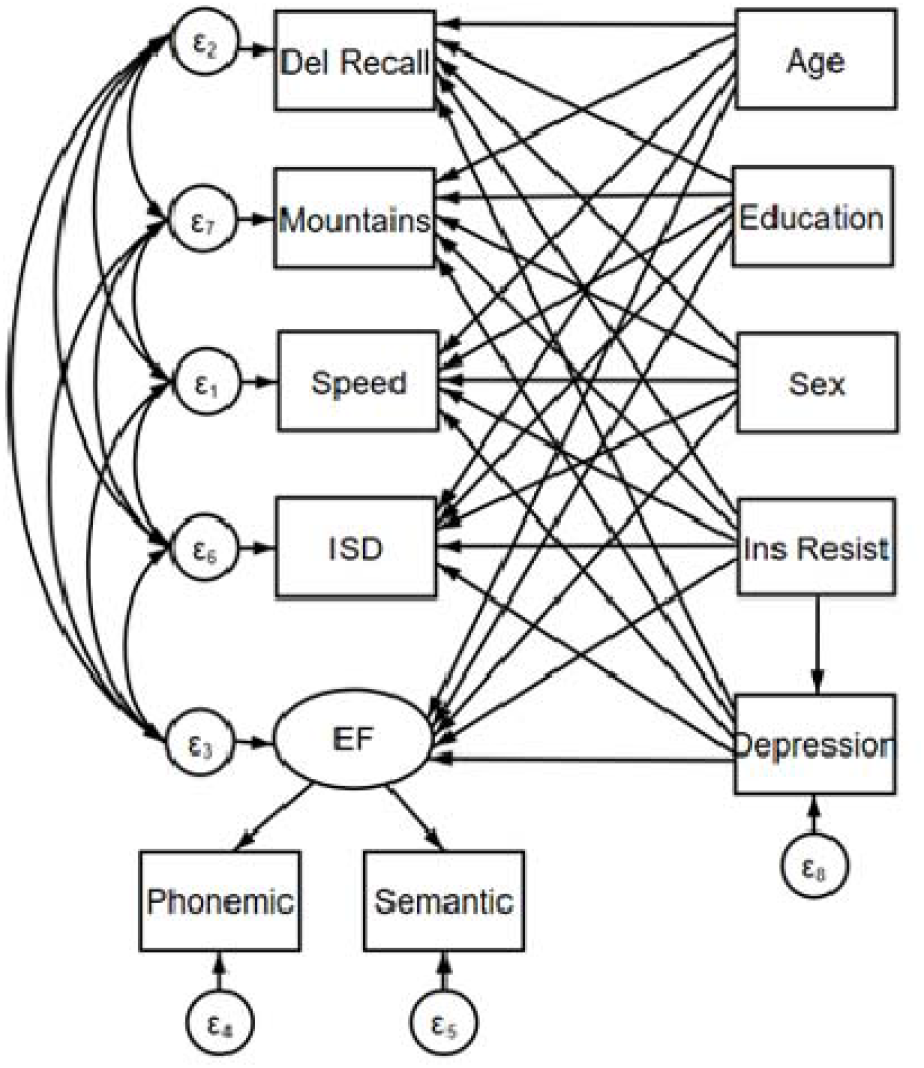
Structural Equation Model Path Diagram

#### 3.3.5 Estimation and Fit

The model was estimated using the maximum likelihood estimation method with missing values (mlmv) method with standardised beta coefficients. The mlmv method assumes joint normality and that missing values are missing at random. The full SEM output is presented in Table 2 where the beta values are presented as standardised values. The results of the SEM analysis showed that higher insulin resistance values significantly predicted lower executive function performance (b = -.12, p < .01) and higher insulin resistance predicted increased depression (b = .15, p < .001), see Figure 2. Insulin resistance was not associated with performance in any other cognitive tasks or ISD but increased depression predicted poorer performance on the 4 Mountains test (b = .14, p < .01). The model was repeated by age group (age 40-49 vs age 50-60) and for the older age group the relationship between lower insulin resistance and executive function remained significant (b = -.15, p < .01) but for the younger age group this relationship was non-significant (b = -.09, p = .126). For both age groups higher insulin resistance predicted increased depression (p < .01), whereas higher depression predicted poorer performance in the 4 Mountains task (p < .05) for only the older age group.

**Table 2:**
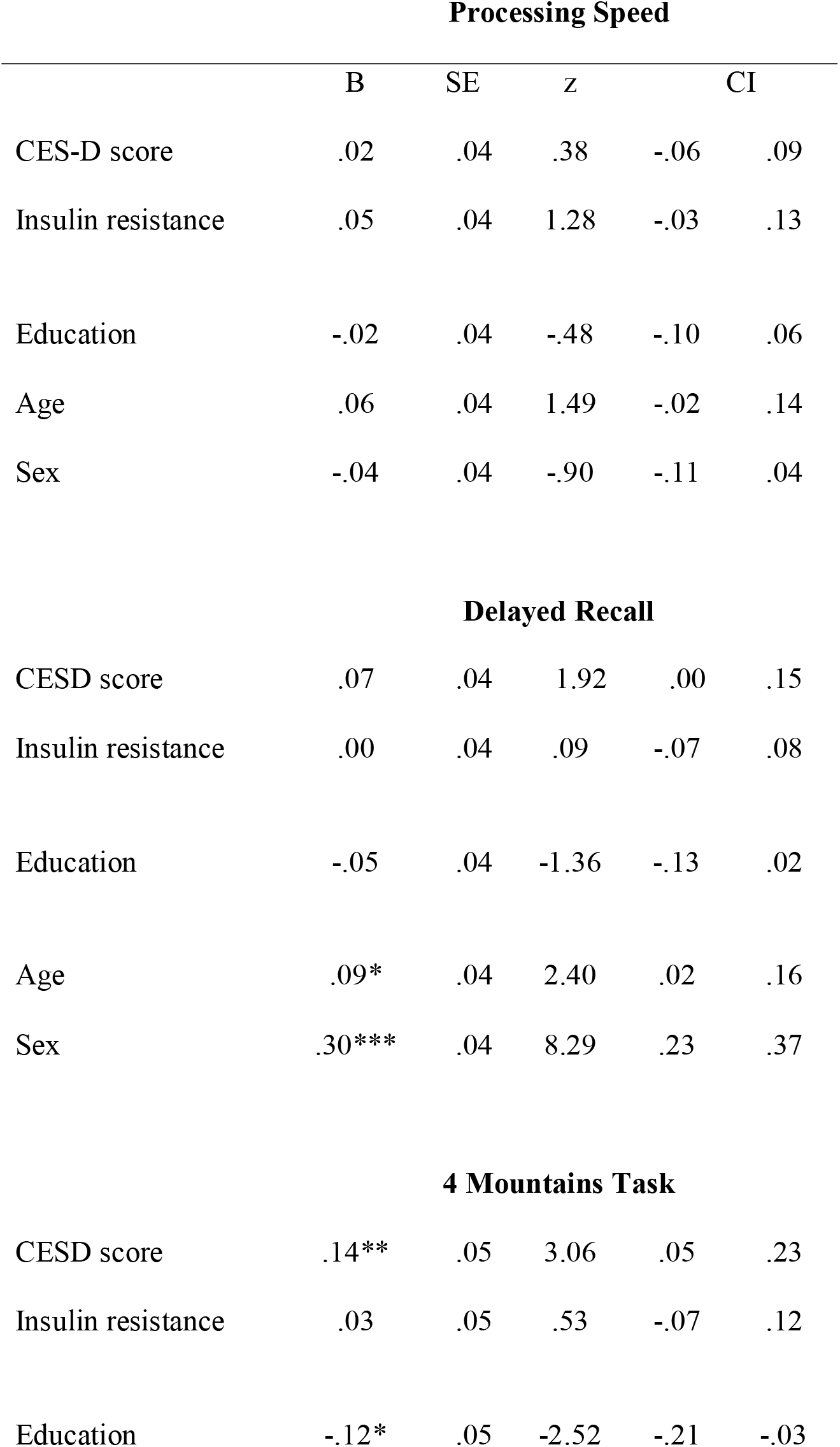
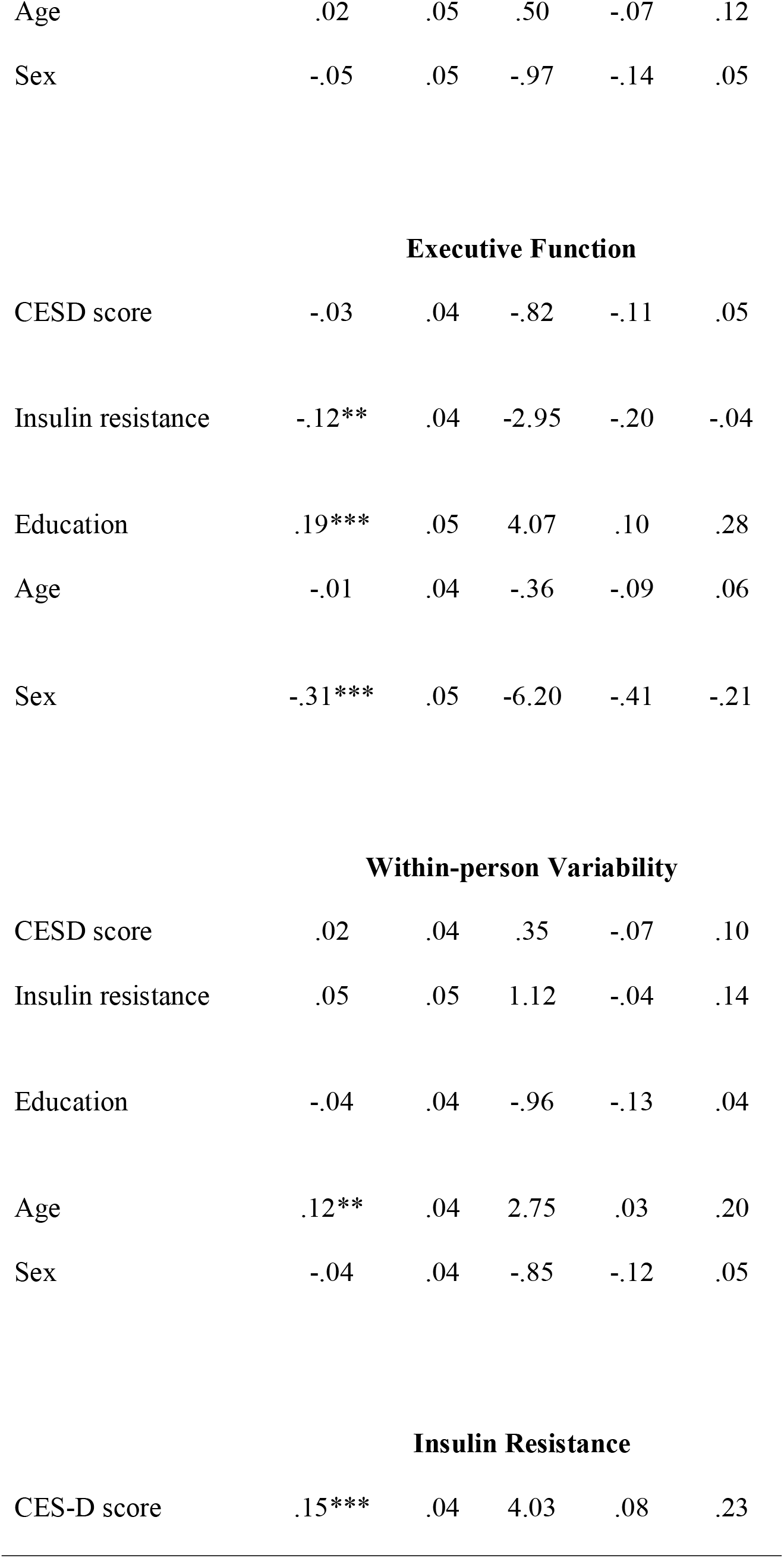
SEM Output.

| **Processing Speed** |  |  |  |  |  |
| --- | --- | --- | --- | --- | --- |
|  | B | SE | z | CI |  |
| CES-D score | .02 | .04 | .38 | -.06 | .09 |
| Insulin resistance | .05 | .04 | 1.28 | -.03 | .13 |
| Education | -.02 | .04 | -.48 | -.10 | .06 |
| Age | .06 | .04 | 1.49 | -.02 | .14 |
| Sex | -.04 | .04 | -.90 | -.11 | .04 |
| **Delayed Recall** |  |  |  |  |  |
| CESD score | .07 | .04 | 1.92 | .00 | .15 |
| Insulin resistance | .00 | .04 | .09 | -.07 | .08 |
| Education | -.05 | .04 | -1.36 | -.13 | .02 |
| Age | .09* | .04 | 2.40 | .02 | .16 |
| Sex | .30*** | .04 | 8.29 | .23 | .37 |
| **4 Mountains Task** |  |  |  |  |  |
| CESD score | .14** | .05 | 3.06 | .05 | .23 |
| Insulin resistance | .03 | .05 | .53 | -.07 | .12 |
| Education | -.12* | .05 | -2.52 | -.21 | -.03 |
| Age | .02 | .05 | .50 | -.07 | .12 |
| Sex | -.05 | .05 | -.97 | -.14 | .05 |
| Executive Function |  |  |  |  |  |
| CESD score | -.03 | .04 | -.82 | -.11 | .05 |
| Insulin resistance | -.12** | .04 | -2.95 | -.20 | -.04 |
| Education | .19*** | .05 | 4.07 | .10 | .28 |
| Age | -.01 | .04 | -.36 | -.09 | .06 |
| Sex | -.31*** | .05 | -6.20 | -.41 | -.21 |
| Within-person Variability |  |  |  |  |  |
| CESD score | .02 | .04 | .35 | -.07 | .10 |
| Insulin resistance | .05 | .05 | 1.12 | -.04 | .14 |
| Education | -.04 | .04 | -.96 | -.13 | .04 |
| Age | .12** | .04 | 2.75 | .03 | .20 |
| Sex | -.04 | .04 | -.85 | -.12 | .05 |
| Insulin Resistance |  |  |  |  |  |
| CES-D score | .15*** | .04 | 4.03 | .08 | .23 |

**Figure 2.**
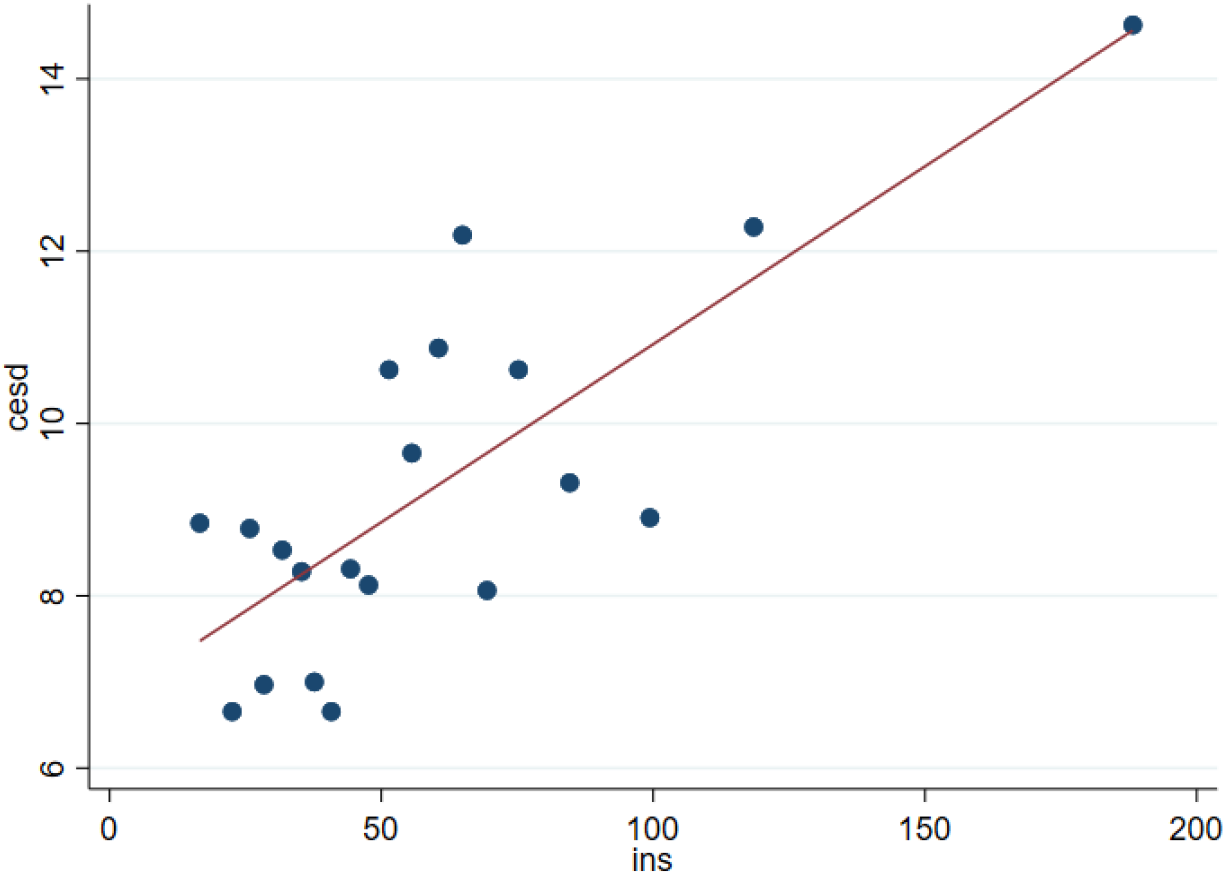
Visualisation of the linear relationship between depression and insulin resistance Note. CESD refers to depression scores and INS refers to insulin resistance as measured by HOMA-IR.

Goodness-of-fit measures for the initial full group model were deemed good by field standards (e.g., Kline, 2016): Root Mean Squared Error of Approximation (RMSEA) = .02; Comparative fit index (CFI) = .983; Tucker Lewis index (TLI) = .925, chi^2^= 15.80, p = .148. RMSEA is a metric of the differences between the predicted outcomes of the model and the observed values in the data. CFI is a metric of the improvement in a model’s fit going from the baseline model to the proposed model, which is less sensitive to differences in sample sizes. TLI is a measure of the relative reduction in misfit per additional degree of freedom in the model.

## 4. Discussion

The aim of the current study was to explore the interrelationships between cognitive functioning, depression, and insulin resistance in cognitively healthy middle-aged adults. Using structural equation modelling, we found that lower insulin resistance values predicted higher executive function performance, whilst controlling for the effects of age, education, and gender. We also found that higher insulin resistance values predicted increased self-reported depression scores. When we re-estimated our models across different age groups (here, 40-49 year olds and 50-60 year olds), we found that the significant relationship between insulin resistance and executive function was apparent for the older age group but not for the younger age group, suggesting that higher insulin resistance in older middle-aged adults is specifically detrimental to aspects of cognition, including executive function. As for the age-related effects on the relationship between insulin resistance and depression, our results revealed that higher levels of insulin resistance were associated with higher levels of depressive symptoms in both age groups. And finally, we found that higher levels of depression predicted lower scores of visuospatial navigation skills (or spatial memory) in the older group but not the younger group of middle-aged adults.

Concerning the negative relationship between insulin resistance and executive function, our results corroborate a growing body of literature, suggesting that those with higher levels of insulin resistance have greater executive dysfunction (e.g., Crochiere et al., 2019). Looking more broadly at available cross-sectional studies assessing the relationship between T2DM and cognitive impairment, many have found that those with T2DM perform more poorly across all cognitive domains, with impairments being most notable in aspects of executive function (e.g., van den Berg et al., 2009). A systematic review of the T2DM-cognition literature found that the average effect size of cognitive impairment in TD2M patients compared to healthy controls was 0.4, which typically represents a moderate effect size (van den Berg et al., 2009). However, in our analysis, we observed that age moderates the relationship between insulin resistance and cognition, which disagrees with the findings of other papers (e.g., Yeung et al., 2009). Given that executive functions are not a unified and homogenous set of neuropsychological constructs, it is possible that the moderating effect of age on the relationship between insulin resistance and executive function holds only for some subtypes of executive functioning but not for others, such as shifting and inhibition in Yeung et al., 2009. Secondly, whilst we used a continuous measure of insulin resistance as a possible indicator of diabetes risk, Yeung and colleagues (2009) used a categorical measure of diabetic status in which only 41 participants of the 465 included in their sample had T2DM. Such use of T2DM caseness as a proxy of insulin resistance is likely under-powered to detect relationships between insulin resistance and cognitive outcomes.

Turning now to our findings of a relationship between greater levels of insulin resistance and higher scores of self-reported depression, in both younger and older adults, the results of the current study fit squarely within the state of the current literature. Similar to our findings, it has been shown that depression is highly comorbid in T2DM with some studies suggesting that depression is two times more common in T2DM compared to the general population (Roy & Lloyd, 2012). Indeed, it has also been shown that depression is related to insulin resistance, which is characterised as a prodromal stage of T2DM in which people with depression and insulin resistance have up to a 60% higher risk of developing clinically diagnosed T2DM in the future (Mezuk et al., 2008). Corroborating our findings of a positive relationship between insulin resistance and depression is the results of a meta-analysis of 18 papers conducted in 2013 by Kan and colleagues. Of the reviewed studies, it was found that the effect size between insulin resistance and depression was estimated to be approximately 0.19, aligning with the results of the current paper. Taken together, it seems that there is growing evidence of a link between chronically low mood and insensitivity to insulin and possible T2DM, which warrants further explanation considering the fact that depression and diabetes are some of the world’s leading causes of socioeconomic burden, decreased life expectancy, and poorer quality of living.

Indeed, given that insulin resistance and depression do seem to be comorbid, theorists have proposed several pathways to understand the link between depression and insensitivity to insulin. In recent years depression has been further characterised as a (partly) inflammatory disorder owning to the potential role of psychosocial and environmental stress in triggering depressive episodes. In response to significant stressors, the body produces higher levels of cortisol that, although inflammatory, are thought to support acute fight or flight responses (Bozovic et al., 2013). With chronic stress and persistently elevated levels of stress hormones in the bloodstream, there can be metabolic dysfunction of carbohydrates in the body (Katsu & Baker, 2021). Thus, stress-induced hypercortisolaemia can result in elevated levels of glucose, which is a major pathway in the development of T2DM. In healthy populations, elevated levels of glucose elicit insulin from the pancreas that facilitates the transport of energy-rich glucose to the body’s vital organs, including the brain. With insulin resistance that may be precipitated by higher levels of stress and inflammatory hormones, metabolic processes in the brain may be hampered, which has been associated with both T2DM and depression in other research (Garcia-Serrano & Duarte, 2020; Su et al., 2014).

Taken together, the results of our study have demonstrated relationships between insulin resistance, as a marker of prediabetes, cognitive dysfunction, and depression in midlife participants. There has been a small, but growing, body of research suggesting that the presence of comorbid diabetes and depression may have an additive effect on cognitive decline in people at risk for dementia. A study by Demakakos and colleagues (2017) showed that in mid-life individuals T2DM and depression were associated with faster decline in distinct areas of cognitive functioning. In a sample of T2DM patients, depression was shown to moderate the rate of cognitive decline to the effect that individuals with comorbid depression had steeper decline trajectories compared to those without comorbid depression (Sullivan et al., 2013). As for depression, it has been shown that psychopathologies which emerge in later life are associated with poorer cognitive outcomes (Bauermeister & Bunce, 2015). Given the limit on attentional resources, it is possible that depression-related biases in attention result in poorer cognitive processing in which there are fewer remaining cognitive resources to support healthy cognitive functioning (Everaert et al., 2014). Given that executive functioning is required to manage chronic conditions like depression and T2DM, it is possible that any deficits in planning and behaviour monitoring may worsen conditions, leading to accelerated neurodegeneration.

Several limitations of the currently reported findings exist. First, while the observed association of executive dysfunction with insulin resistance points to likely cerebrovascular pathophysiology, the study lacks biomarker data relevant to dementia to clarify the nature of any potential neurodegenerative processes. Second, while we chose a well-validated measure of insulin resistance in HOMA-IR, the gold standard remains the hyperinsulinemic euglycemic clamp (HEC). It is therefore possible that the reliability of the insulin resistance scores could have been improved by the use of an HEC. The third methodological caveat is that it is now recognised that there is a distinction between central and peripheral insulin resistance which has relevance to human behaviour. HOMA-IR as well as HEC only allow assessment of peripheral insulin resistance; it is possible that the link between depression, insulin resistance and cognition could be clarified further through methods of assessing central insulin resistance specifically.

## 5. Conclusions

Given the evident comorbidities, the current study sought to further explore the interrelationships between cognitive function, depression, and insulin resistance. Using data from over 600 participants from the PREVENT prospective cohort study, our analyses revealed that insulin resistance predicts executive dysfunction in older but not younger middle-aged adults, in addition to scores of depression in both age groups. As for cognitive functioning, depression could predict visuospatial navigation abilities, as measured by a 4 Mountains task, in the older but not younger middle-aged adult group. Together, we have shown connections between three common diseases that place an emotional, health, and socioeconomic burden on individuals and on society at large. It is hoped that the results of the current study will encourage further research into the interrelationships between these diseases with the aim of identifying disease modifying treatments, risk pathways between the disorders, and critical points of intervention to alleviate the undesirable sequalae of being diagnosed with one or multiple commodities.

## Data Availability

All data produced in the present study are available upon reasonable request to the authors

## Acknowledgements

The authors would like to acknowledge and kindly thank the PREVENT study group and the volunteers that contributed to the study.

## CRediT authorship contribution statement

**Sarah Bauermeister:** Conceptualisation, Data Curation, Formal Analysis, Visualization, Writing – Original Draft, and Writing – Review & Editing. **Graham Reid:** Writing – Original Draft, Writing – Review & Editing, and Validation. **Michael Ben Yehuda**: Data Curation, Formal Analysis, and Writing – Review & Editing. **Gregory Howgego:** Writing – Review & Editing. **Karen Ritchie:** Methodology, Supervision, and Writing – Review & Editing. **Tam Watermeyer:** Conceptualisation, Methodology, and Writing – Reviewing & Editing. **Sarah Gregory**: Conceptualisation, Methodology, and Writing – Reviewing & Editing. **Graciela Muniz Terrera**: Methodology, Supervision, Writing – Review & Editing. **Ivan Koychev:** Conceptualisation, Methodology, Writing – Original Draft, Writing – Review & Editing, Supervision, and Project Administration.

