## Supplemental Table 1 for "Insulin Resistance’s Impact on Cognition in Middle Aged Adults from the PREVENT cohort: Interactive Effects with Depression"

**Supplementary Material 1**

Bivariate Correlation Table

|  | Age | Sex | EDUC | IR | CESD | DR | MOUNT | RT | VF PH | VF SEM |
| --- | --- | --- | --- | --- | --- | --- | --- | --- | --- | --- |
| Age |  |  |  |  |  |  |  |  |  |  |
| Sex | .06 |  |  |  |  |  |  |  |  |  |
| EDUC | -.11 | -.08 |  |  |  |  |  |  |  |  |
| IR | .12 | .16* | -.10 |  |  |  |  |  |  |  |
| CESD | -.004 | -.02 | -.04 | .15* |  |  |  |  |  |  |
| DR | .11 | .31* | -.09 | .07 | .07 |  |  |  |  |  |
| MOUNT | .03 | -.02 | -.11 | .05 | .15 | .10 |  |  |  |  |
| RT | .06 | -.02 | -.03 | .05 | .02 | -.02 | .04 |  |  |  |
| VF PH | -.06 | -.06 | .11 | -.04 | -.05 | -.17* | -.06 | .006 |  |  |
| VF SEM | -.05 | -.33* | .22* | -.19* | -.05 | -.22* | -.10 | -.05 | .30* |  |

Abbreviations: EDUC (Education in years); IR (Insulin Resistance); CESD (Epidemiological Studies-Depression Scale); MOUNT (4 Mountains Task); RT (Reaction Time); VF PH (Phonemic Verbal Fluency); VF SEM (Sematic Verbal Fluency)
